# Adherence to the CONSORT statement and risk of bias assessment in Randomized Controlled Trials in rehabilitation journals: a protocol for a meta-research study

**DOI:** 10.1101/2021.01.21.21250256

**Authors:** Tiziano Innocenti, Silvia Giagio, Stefano Salvioli, Daniel Feller, Silvia Minnucci, Fabrizio Brindisino, Raymond Ostelo, Alessandro Chiarotto

## Abstract

**Objective:** The aim of this study will be to assess the adherence to the reporting quality standards set forth in the CONSORT Statement checklist of a random sample of randomized controlled trials (RCTs) published in rehabilitation journals, and to assess the association between this adherence and the risk of bias of these RCTs.

**Methods and Analysis:** A cross-sectional analysis is planned on a random sample of 200 RCTs published between 2011and 2020 in the 68 journals indexed under “rehabilitation” category in InCites Journal Citation Report.

Randomization will be stratified by publication date and journal ranking (quartile range; Q1-2 and Q3-4) to include an equal number of studies from 2011 to 2015 (Q1-Q2=50 and Q3-Q4=50) and from 2016 to 2020 (Q1-Q2=50 and Q3-Q4=50). RCT with parallel group design will be included. Observational or cohort studies, interim analyses, economic analyses of RCTs, RCT protocols, quasi-experimental design post-trial follow-up studies, subgroup and secondary analyses of previously reported RCTs, RCT with cross-over design, pilot feasibility RCTs, n-of-1 trials, cluster trials, editorials, letters and news reports will be excluded. The primary analysis will address the completeness of the reporting for each study and the relationship between CONSORT adherence and risk of bias. This will be a descriptive analysis through descriptive statistics and graphical representation.

**Ethics and Dissemination:** Several studies have shown the positive influence of reporting guidelines on the completeness of research reporting but no one investigated the use and the appropriateness of reporting guidelines in physical therapy research. Therefore, this study will add relevant knowledge that may contribute to improve further the reporting of rehabilitation research. The results of this research will be published in a peer-reviewed journal and will be presented at relevant (inter)national scientific events.

## INTRODUCTION

Research in health sciences should have the potential to advance scientific understanding, or to improve the treatment or prevention of disease. Clinical practice and public health policy decisions should be based on high-quality research findings^1^. The purpose of a research report (e.g. a scientific article) is to communicate the design, execution, and findings of a study with precision and accuracy^2^; this is relevant for several stakeholder groups, including researchers, clinicians, policy makers and patients^3^. To be useful to all these categories, research reports should include all relevant information about methods and results. Additionally, accurate reporting of a study is essential to judge its validity and the clinical applicability of its findings^1^.

Problems about reporting of a scientific study can affect research in different ways. For example, it is known that study methods are frequently not described in adequate detail and that results are presented ambiguously, incompletely or selectively^3^. The consequence is that many reports cannot be used for replication studies, or they are even harmful, as well as a waste of resources^4^. Moreover, the validity is hard to judge and therefore these studies are difficult to use for decision making in health care/clinical guidelines. To overcome these problems, reporting guidelines (RGs) have been developed to support authors in reporting research methods and results. In 2014, 28 rehabilitation journals have simultaneously published an editorial to highlight the need of using RGs to ensure the quality and the completeness of studies in the field^5^. Chan and colleagues concluded the editorial hope: “that simultaneous implementation of this new reporting requirement will send a strong message to all disability and rehabilitation researchers about the need to adhere to the highest standards when performing and disseminating research.”

When assessing the effectiveness of an intervention for a clinical condition, randomized controlled trials (RCTs) are considered the preferable source of evidence^6^. Assessing the validity of a trial is dependent on the completeness and transparency of the study report^6^. This requires the inclusion of key study information so that readers can properly assess the validity and generalizability of each study and apply the findings to their patient population. Responsibility extends to peer reviewers and medical journal editors who must verify that the information needed to evaluate study quality is reported in manuscripts, and some guidelines exist to ensure that the essential elements are reported in the manuscript^7^. The CONSORT (Consolidated Standards of Reporting Trials) statement is used worldwide as a reporting guideline focused on RCTs^8^. The statement was published in 1996^9^, updated in 2010^10^, and consists of 25 key items checklist (with 11 sub-item, for a total of 37 items) that guide the reporting of an RCT. Assessing published trials for their completeness (i.e., adherence to CONSORT checklists) is important for directing further publication policies and for minimizing the risk of selective and/or publication bias^11^.

Critical appraisal of study methods enables judgements of the degree to which results and author interpretations overestimate or under-estimate study effects, and is highly dependent on what is reported within research reports. For example, if a study has better reporting, this could make more adequately the evaluation of risk of bias^12^. The tool most frequently used to assess the risk of bias of RCTs is the Cochrane risk of bias tool^13^. A revised version of the tool (ROB 2) was published in 2019^14^, it is structured in five domains (all mandatory, and none can be added) and an overall risk of bias judgment. The risk of bias judgment for each domain can be expressed as ‘‘low’’, ‘‘high’’ or ‘‘some concern’’, and the overall risk of bias normally corresponds to the lowest judgment in any of the domains. This assessment should be made for each outcome measure considered critical or important.

Many previous studies assessed the adherence to CONSORT checklist in RCTs in the medical field^15, 16^. These studies have shown poor overall adherence to the CONSORT checklist items, especially for the items related to the methods section. Another previous study^17^ confirmed that most of the authors (∼ 88%) publishing RCTs in high impact rehabilitation journals did not mention RG use and approximately half of those who declared using RGs did not do it in an appropriate manner. This occurred despite many rehabilitation journals requiring RGs as mandatory during submission. To our knowledge, the adherence to the CONSORT checklist and the relationship between risk of bias and the completeness of the reporting in RCTs of rehabilitation interventions has not been systematically evaluated.

The aim of this study will be to assess the adherence to the reporting quality standards set forth in the CONSORT Statement checklist of a random sample of RCTs published in rehabilitation journals, and to assess the association between this adherence and the risk of bias of these RCTs.

### Objectives

#### Primary Objectives

1. To evaluate the completeness of the reporting in RCTs published in rehabilitation journals between 2011 and 2020 by adherence to indications contained in the CONSORT Checklist^10^.
2. To investigate the possible relationship between completeness of reporting and risk of bias.

### Secondary Objectives

1) To compare the adherence to the CONSORT Checklist^10^ and year of publication, journal proprieties (e.g. Quartile, Open Access/Subscription) and others factors (e.g. registration of the study protocol) in order to investigate the impact of these features.

## METHODS AND ANALYSIS

We will conduct a cross-sectional analysis in a random sample of 200 RCTs published between 2011and 2020 in the 68 journals indexed under “rehabilitation” category in InCites Journal Citation Report^18^. This sampling was chosen to include all the journals in the rehabilitation field. The 68 rehabilitation journals indexed in InCites are reported in Table 1.

**Table 1:**
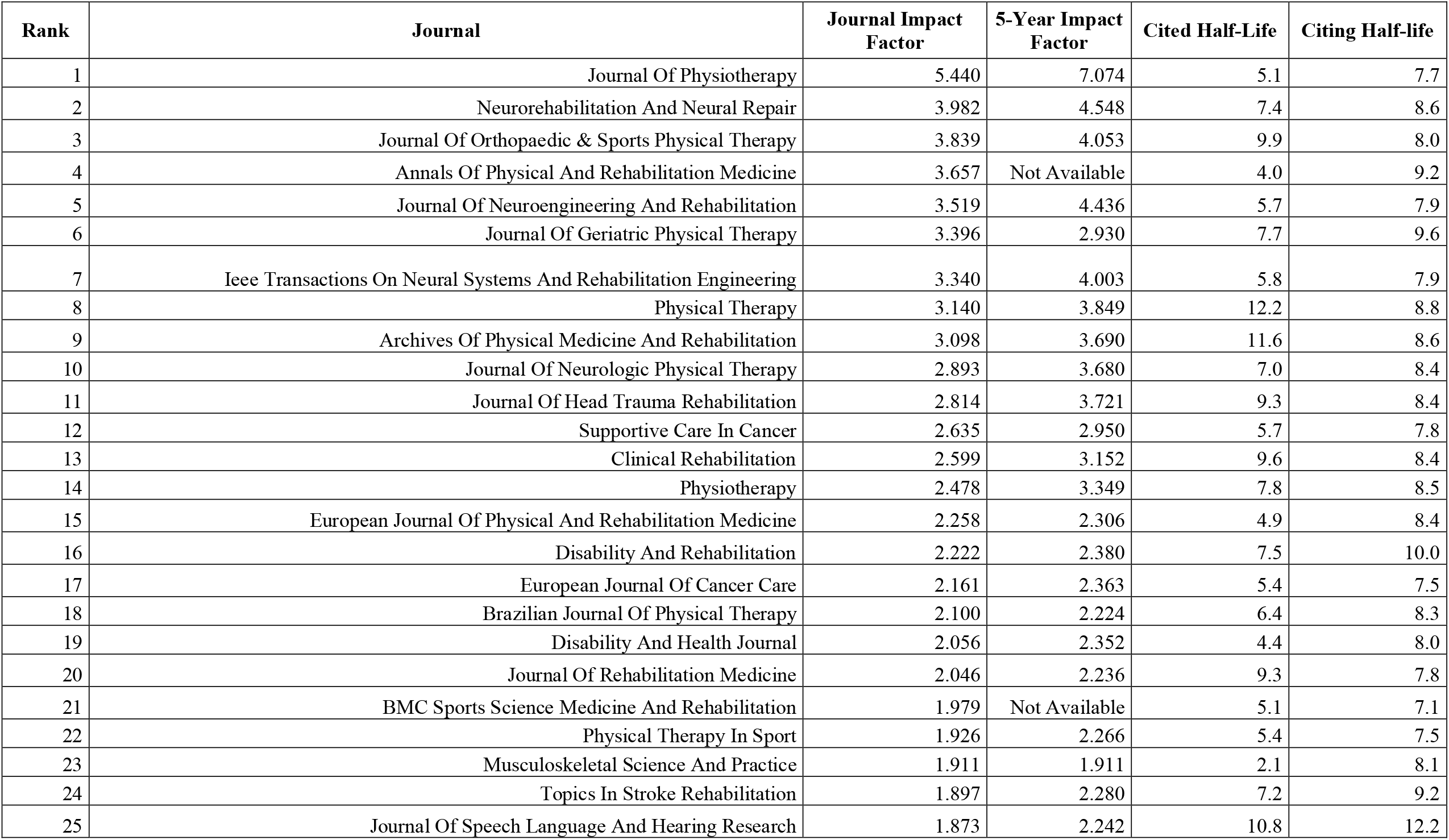
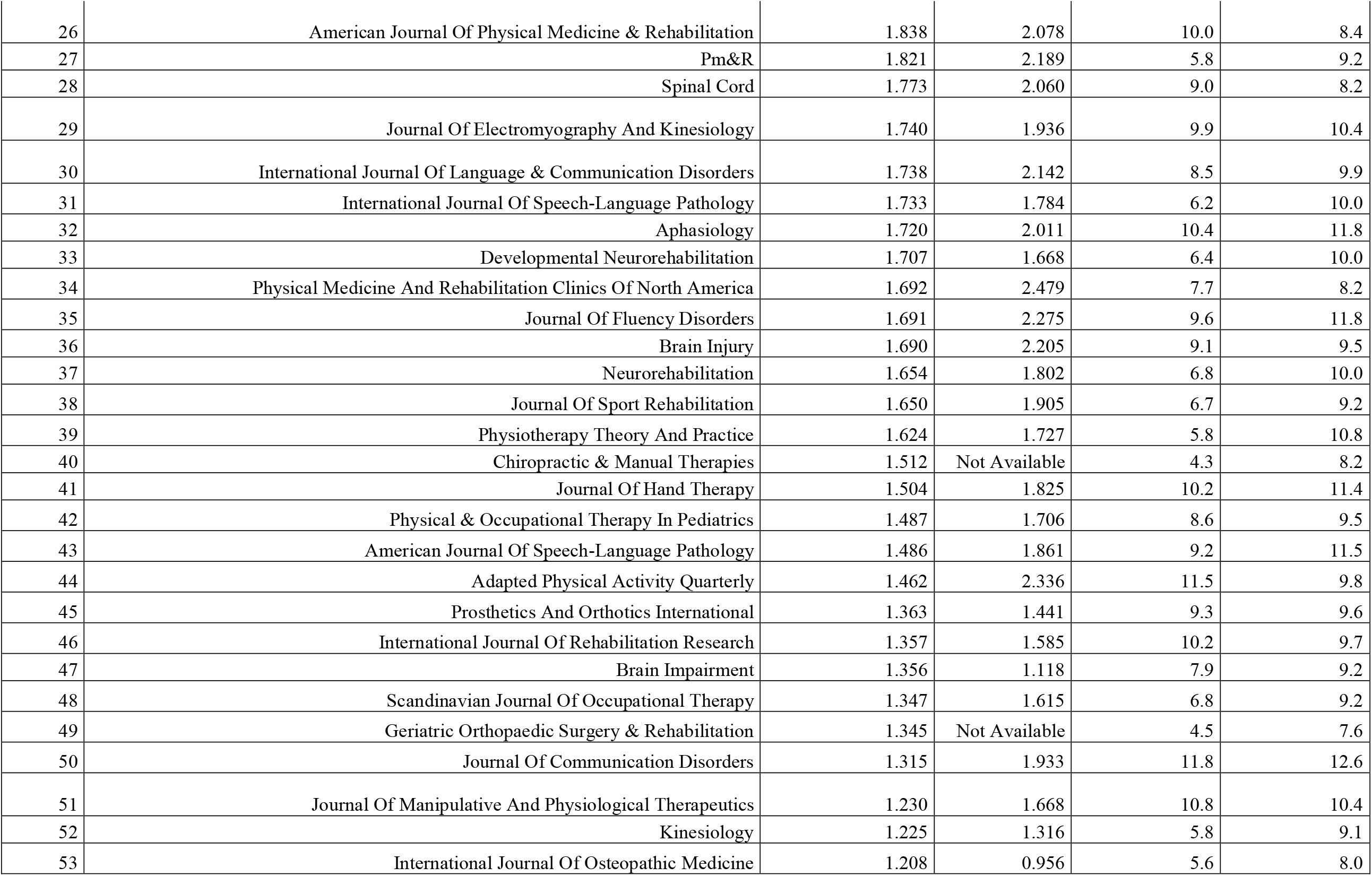
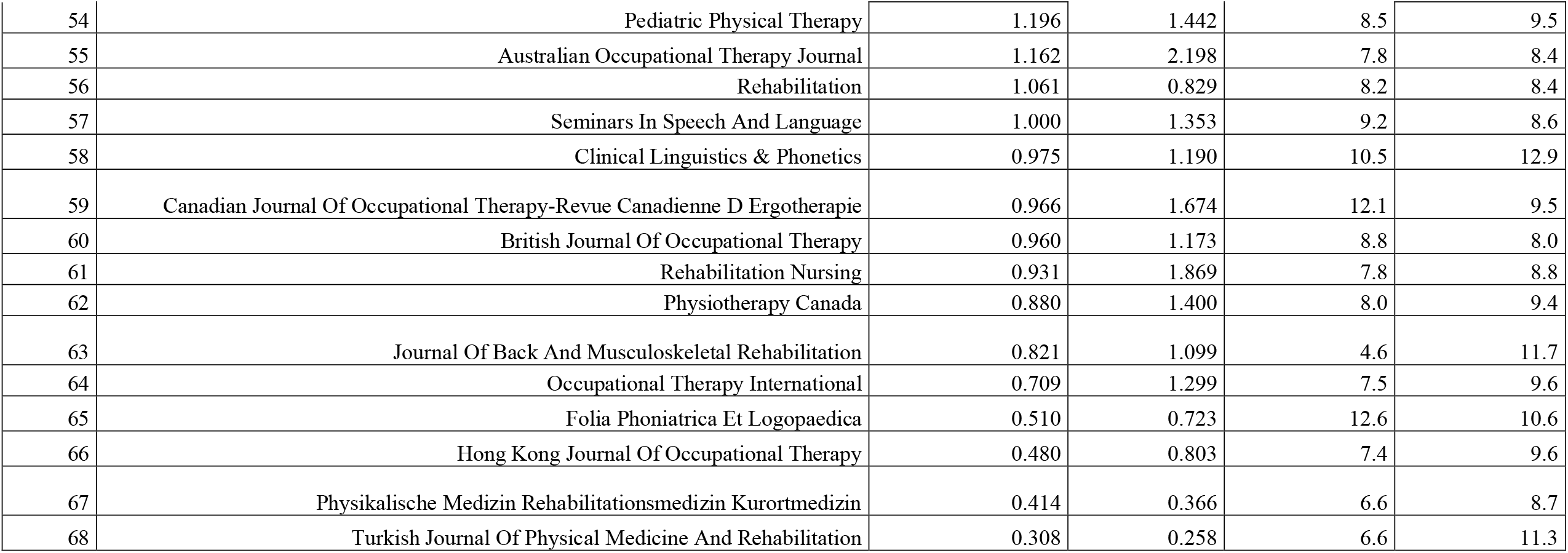
Journals selected (Year 2019 according InCites Journal Citation Reports)

| Rank | Journal | Journal Impact Factor | 5-Year Impact Factor | Cited Half-Life | Citing Half-life |
| --- | --- | --- | --- | --- | --- |
| 1 | Journal Of Physiotherapy | 5.440 | 7.074 | 5.1 | 7.7 |
| 2 | Neurorehabilitation And Neural Repair | 3.982 | 4.548 | 7.4 | 8.6 |
| 3 | Journal Of Orthopaedic & Sports Physical Therapy | 3.839 | 4.053 | 9.9 | 8.0 |
| 4 | Annals Of Physical And Rehabilitation Medicine | 3.657 | Not Available | 4.0 | 9.2 |
| 5 | Journal Of Neuroengineering And Rehabilitation | 3.519 | 4.436 | 5.7 | 7.9 |
| 6 | Journal Of Geriatric Physical Therapy | 3.396 | 2.930 | 7.7 | 9.6 |
| 7 | Ieee Transactions On Neural Systems And Rehabilitation Engineering | 3.340 | 4.003 | 5.8 | 7.9 |
| 8 | Physical Therapy | 3.140 | 3.849 | 12.2 | 8.8 |
| 9 | Archives Of Physical Medicine And Rehabilitation | 3.098 | 3.690 | 11.6 | 8.6 |
| 10 | Journal Of Neurologic Physical Therapy | 2.893 | 3.680 | 7.0 | 8.4 |
| 11 | Journal Of Head Trauma Rehabilitation | 2.814 | 3.721 | 9.3 | 8.4 |
| 12 | Supportive Care In Cancer | 2.635 | 2.950 | 5.7 | 7.8 |
| 13 | Clinical Rehabilitation | 2.599 | 3.152 | 9.6 | 8.4 |
| 14 | Physiotherapy | 2.478 | 3.349 | 7.8 | 8.5 |
| 15 | European Journal Of Physical And Rehabilitation Medicine | 2.258 | 2.306 | 4.9 | 8.4 |
| 16 | Disability And Rehabilitation | 2.222 | 2.380 | 7.5 | 10.0 |
| 17 | European Journal Of Cancer Care | 2.161 | 2.363 | 5.4 | 7.5 |
| 18 | Brazilian Journal Of Physical Therapy | 2.100 | 2.224 | 6.4 | 8.3 |
| 19 | Disability And Health Journal | 2.056 | 2.352 | 4.4 | 8.0 |
| 20 | Journal Of Rehabilitation Medicine | 2.046 | 2.236 | 9.3 | 7.8 |
| 21 | BMC Sports Science Medicine And Rehabilitation | 1.979 | Not Available | 5.1 | 7.1 |
| 22 | Physical Therapy In Sport | 1.926 | 2.266 | 5.4 | 7.5 |
| 23 | Musculoskeletal Science And Practice | 1.911 | 1.911 | 2.1 | 8.1 |
| 24 | Topics In Stroke Rehabilitation | 1.897 | 2.280 | 7.2 | 9.2 |
| 25 | Journal Of Speech Language And Hearing Research | 1.873 | 2.242 | 10.8 | 12.2 |
| 26 | American Journal Of Physical Medicine & Rehabilitation | 1.838 | 2.078 | 10.0 | 8.4 |
| 27 | Pm&R | 1.821 | 2.189 | 5.8 | 9.2 |
| 28 | Spinal Cord | 1.773 | 2.060 | 9.0 | 8.2 |
| 29 | Journal Of Electromyography And Kinesiology | 1.740 | 1.936 | 9.9 | 10.4 |
| 30 | International Journal Of Language & Communication Disorders | 1.738 | 2.142 | 8.5 | 9.9 |
| 31 | International Journal Of Speech-Language Pathology | 1.733 | 1.784 | 6.2 | 10.0 |
| 32 | Aphasiology | 1.720 | 2.011 | 10.4 | 11.8 |
| 33 | Developmental Neurorehabilitation | 1.707 | 1.668 | 6.4 | 10.0 |
| 34 | Physical Medicine And Rehabilitation Clinics Of North America | 1.692 | 2.479 | 7.7 | 8.2 |
| 35 | Journal Of Fluency Disorders | 1.691 | 2.275 | 9.6 | 11.8 |
| 36 | Brain Injury | 1.690 | 2.205 | 9.1 | 9.5 |
| 37 | Neurorehabilitation | 1.654 | 1.802 | 6.8 | 10.0 |
| 38 | Journal Of Sport Rehabilitation | 1.650 | 1.905 | 6.7 | 9.2 |
| 39 | Physiotherapy Theory And Practice | 1.624 | 1.727 | 5.8 | 10.8 |
| 40 | Chiropractic & Manual Therapies | 1.512 | Not Available | 4.3 | 8.2 |
| 41 | Journal Of Hand Therapy | 1.504 | 1.825 | 10.2 | 11.4 |
| 42 | Physical & Occupational Therapy In Pediatrics | 1.487 | 1.706 | 8.6 | 9.5 |
| 43 | American Journal Of Speech-Language Pathology | 1.486 | 1.861 | 9.2 | 11.5 |
| 44 | Adapted Physical Activity Quarterly | 1.462 | 2.336 | 11.5 | 9.8 |
| 45 | Prosthetics And Orthotics International | 1.363 | 1.441 | 9.3 | 9.6 |
| 46 | International Journal Of Rehabilitation Research | 1.357 | 1.585 | 10.2 | 9.7 |
| 47 | Brain Impairment | 1.356 | 1.118 | 7.9 | 9.2 |
| 48 | Scandinavian Journal Of Occupational Therapy | 1.347 | 1.615 | 6.8 | 9.2 |
| 49 | Geriatric Orthopaedic Surgery & Rehabilitation | 1.345 | Not Available | 4.5 | 7.6 |
| 50 | Journal Of Communication Disorders | 1.315 | 1.933 | 11.8 | 12.6 |
| 51 | Journal Of Manipulative And Physiological Therapeutics | 1.230 | 1.668 | 10.8 | 10.4 |
| 52 | Kinesiology | 1.225 | 1.316 | 5.8 | 9.1 |
| 53 | International Journal Of Osteopathic Medicine | 1.208 | 0.956 | 5.6 | 8.0 |
| 54 | Pediatric Physical Therapy | 1.196 | 1.442 | 8.5 | 9.5 |
| 55 | Australian Occupational Therapy Journal | 1.162 | 2.198 | 7.8 | 8.4 |
| 56 | Rehabilitation | 1.061 | 0.829 | 8.2 | 8.4 |
| 57 | Seminars In Speech And Language | 1.000 | 1.353 | 9.2 | 8.6 |
| 58 | Clinical Linguistics & Phonetics | 0.975 | 1.190 | 10.5 | 12.9 |
| 59 | Canadian Journal Of Occupational Therapy-Revue Canadienne D Ergotherapie | 0.966 | 1.674 | 12.1 | 9.5 |
| 60 | British Journal Of Occupational Therapy | 0.960 | 1.173 | 8.8 | 8.0 |
| 61 | Rehabilitation Nursing | 0.931 | 1.869 | 7.8 | 8.8 |
| 62 | Physiotherapy Canada | 0.880 | 1.400 | 8.0 | 9.4 |
| 63 | Journal Of Back And Musculoskeletal Rehabilitation | 0.821 | 1.099 | 4.6 | 11.7 |
| 64 | Occupational Therapy International | 0.709 | 1.299 | 7.5 | 9.6 |
| 65 | Folia Phoniatica Et Logopaedica | 0.510 | 0.723 | 12.6 | 10.6 |
| 66 | Hong Kong Journal Of Occupational Therapy | 0.480 | 0.803 | 7.4 | 9.6 |
| 67 | Physikalische Medizin Rehabilitationsmedizin Kurortmedizin | 0.414 | 0.366 | 6.6 | 8.7 |
| 68 | Turkish Journal Of Physical Medicine And Rehabilitation | 0.308 | 0.258 | 6.6 | 11.3 |

### Study selection criteria

RCT with parallel group design published from 2011 to 2020 as full-text scientific articles will be included. Observational or cohort studies, interim analyses, economic analyses of RCTs, RCT protocols, quasi-experimental design post-trial follow-up studies, subgroup and secondary analyses of previously reported RCTs, RCT with cross-over design, pilot feasibility RCTs, n-of-1 trials, cluster trials, editorials, letters and news reports will be excluded.

### Study selection process

Journal tags for the journals will be identified in Medline and a detailed search strategy will be created to find all RCTs published from 2011 to 2020 in this database (See Table 2). The searching will be performed only in Medline because all of the journals are indexed in Medline.

**Table 2:**
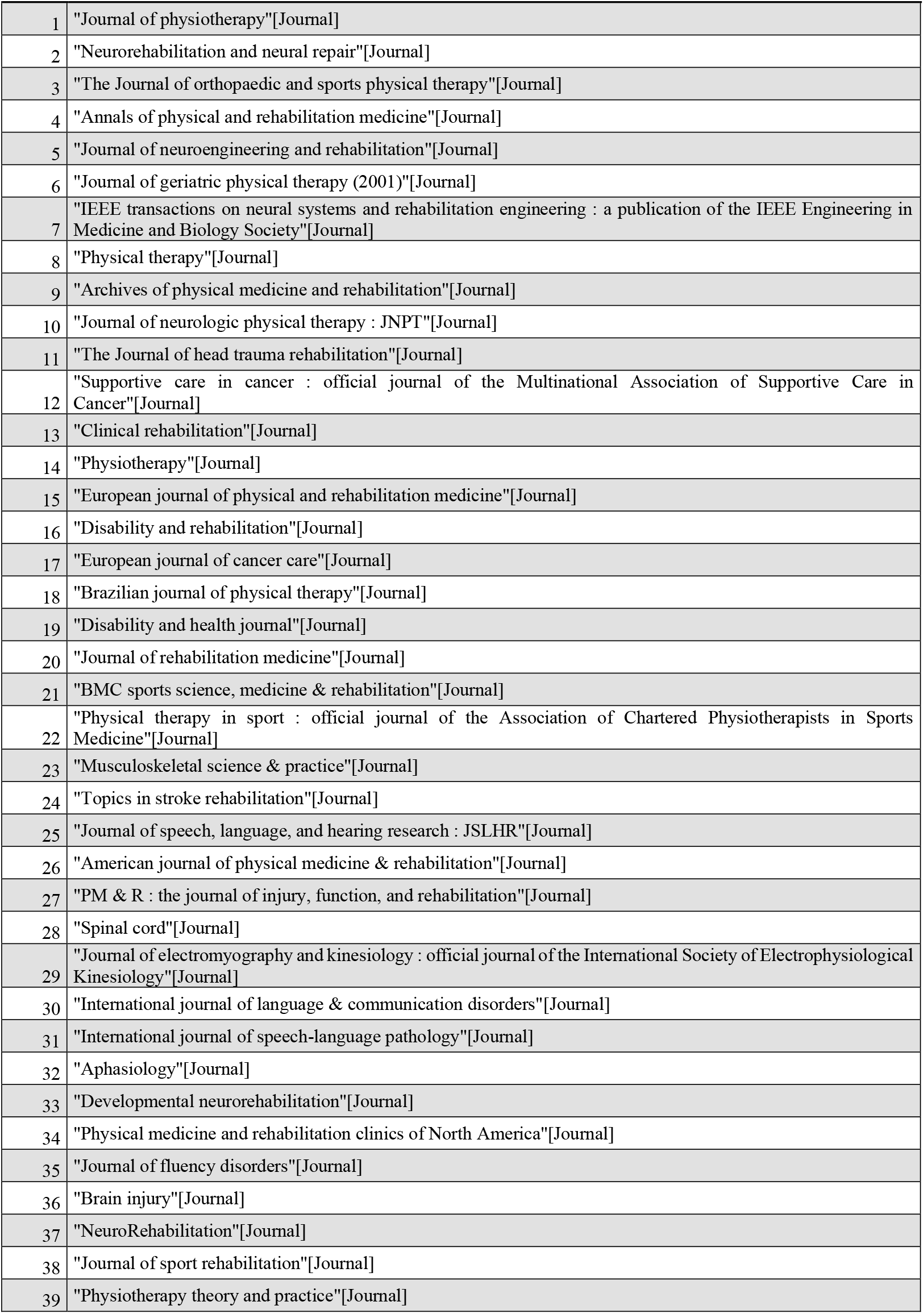
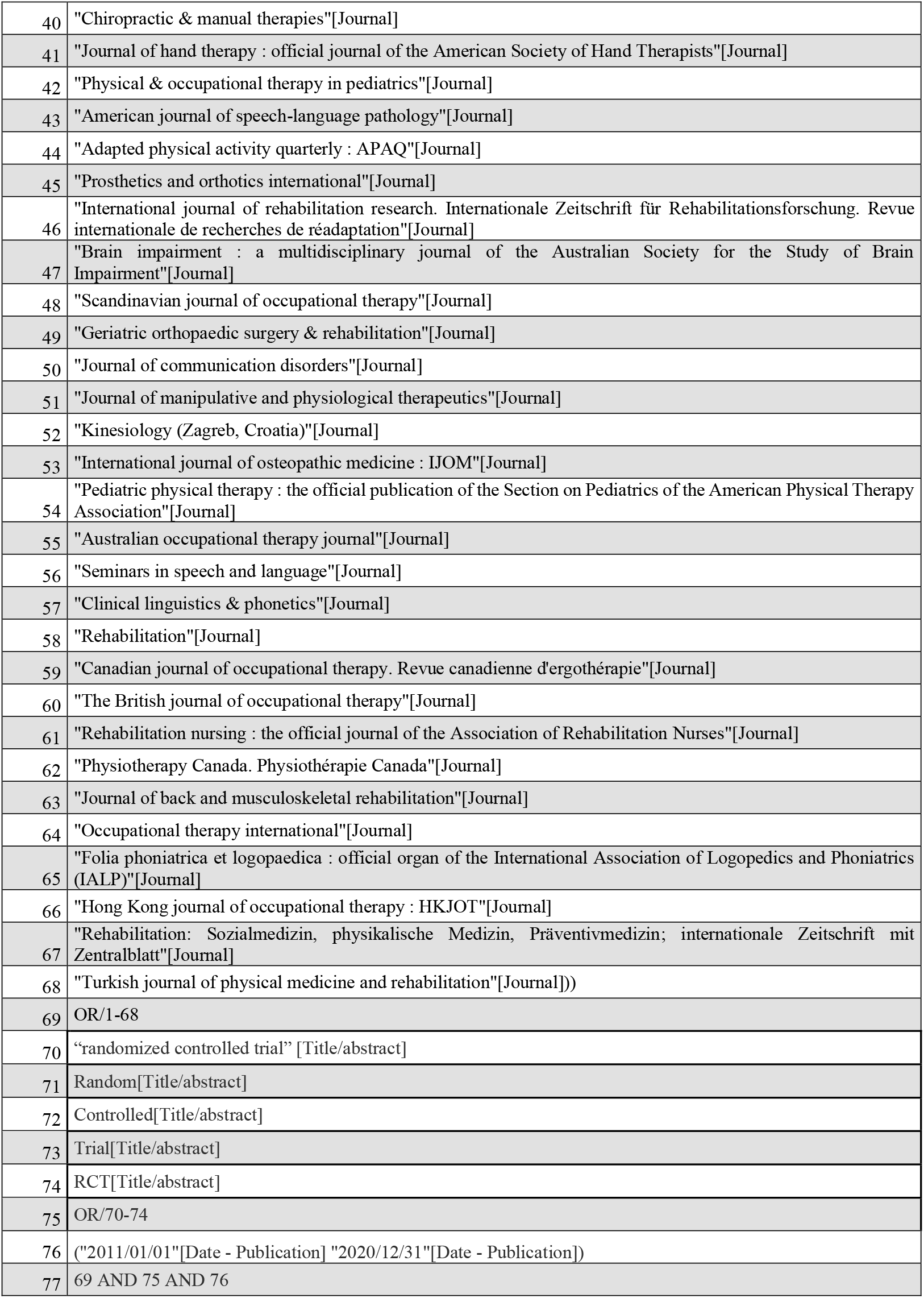
Search Strategy.

Titles and abstracts of the selected articles will be screened for eligibility in a double blinded process by two independent reviewers who select potentially eligible articles independently and based on selection criteria. Any disagreement will be resolved by a third reviewer. Subsequently, the study sample of 200 hits will be selected using a computerized random sequence generator^19^. Randomization will be stratified by publication date and journal ranking (quartile range; Q1-2 and Q3-4) to include an equal number of studies from 2011 to 2015 (Q1-Q2=50 and Q3-Q4=50) and from 2016 to 2020 (Q1-Q2=50 and Q3-Q4=50). Study selection process will be summarized in a tailored flow diagram.

### Data extraction

Full-texts will be stored in EndNote X7 (Thomas Reuters, Philadelphia, Pennsylvania, USA). A data extraction form will be used by two reviewers and the following data will be extracted:

- First author and year of publication;
- Journal with characteristic (Quartile Range, Publication options - Open Access/subscription/Hybrid – mandatory use of RGs);
- Country in which was conducted;
- Rehabilitation research field (e.g. musculoskeletal, neurological, pediatrics, pelvic-floor disfunction, cardio-pulmonary, other)
- Published protocol (Yes/No)
- Adherence to the CONSORT checklist (see later);
- Risk of bias with the Cochrane RoB 2 checklist (see later).

Data extraction and the risk of bias assessment from each RCT will be performed by two reviewers, independently; when necessary, a third researcher will solve disagreements.

### Assessing the completeness of the reporting

We will determine compliance to each item of the CONSORT checklist through the use of 25 item checklist. Discrepancies between authors over application of checklist items will be resolved by consulting the published explanation of the CONSORT checklist^10^. Prior to scoring of study inter-rater agreement for each item on the 25-item checklist will be evaluated in a pilot study of 20 RCTs and scored independently by two authors with post-graduate training in epidemiology and critical appraisal.

According to the explanation and elaboration statements of CONSORT Checklist, each item will be marked with “1” if it was well described, incomplete or missing items with “0”, and not applicable items with “NA”. Total adherence for each item between studies and total adherence to the CONSORT Checklist for each study will be calculated. Given the aim of this study is to investigate the completeness of the reporting, authors of included studies will be not contacted for information omitted from manuscripts. Summary tables and graphics of extracted data of all included studies and a narrative synthesis will be provided.

### Assessing the risk of bias

RoB 2 tool^14^ analysis will be used to assess the risk of bias in the included studies. Since this assessment should be made for each outcome measure considered critical or important, we will consider the primary outcome(s) of each RCTs. For each RCT a risk of bias judgment will be reached for each domain (bias arising from the randomization process; bias due to deviations from intended interventions; bias due to missing outcome data; bias in measurement of the outcome; and bias in selection of the reported result) as suggested by Cochrane Collaboration^14^. The judgment about risk of bias will be ‘‘high,’’ ‘‘low,’’ or ‘‘some concerns’’. The results will be graphically summarized through the risk of bias graph obtained with the ROBVIS Tool^20^. Risk of bias assessment from each RCT will be performed by two reviewers, independently; when necessary, a third researcher will solve disagreements.

## Data analysis

The primary analysis will address:

- The completeness of the reporting for each study. Total adherence to the CONOSORT checklist will be calculated (in percentage) as the total number of items described and reported out of the total number of applicable items.
- The total adherence for each item between studies, calculated as (for each item; in percentage) the number of times that one item is described and reported out of the total number of studies that have such item applicable.
- The relationship between CONSORT adherence and risk of bias. RoB 2 is not an unidimensional tool, therefore we will not perform a quantitative analysis due to the nature and multidimensionality of both instruments (ROB 2 and CONSORT Checklist). This will be a descriptive analysis through descriptive statistics and graphical representation.

Secondary analysis:

- The impact of time elapsed (per year) since publication of the articles and total adherence will be evaluated. A regression analysis will be performed to assess the relationship between the adherence to the CONSORT checklist and the year of publication, with use of total adherence (as described above) as dependent variable and publication year (2011-2020) as independent variable.
- In the same way, the relationship between the following characteristics (as independent variables) and the total adherence will be evaluated:
  - Journal quartile
  - Publications modalities (e.g. Open Access vs Subscription vs Hybrid)
  - Published study protocol

## Data Availability

All data are reported in the manuscript.

## ETHICS AND DISSEMINATION

Several studies have shown the positive influence of RGs on the completeness of research reporting^11^. The adherence to these guidelines can result in a published article that contains precise information and that can better allow readers to make informed judgments. By increasing the likelihood that critical information is included in the submitted manuscript, the work of editors and external reviewers can also become more efficient^2^.

Since the publication of the multi-journal editorial of Chan and colleagues^5^, no study investigated the adherence to these RGs in physical therapy research.

Therefore, this study will add relevant knowledge that may contribute to improve further the value of rehabilitation research.

This study will also align with the mission statement of the EQUATOR (Enhancing the QUAlity and Transparency of health Research) Network^21^, an international collaboration aiming to enhance the reliability of medical research literature by promoting transparent, accurate reporting of research studies.. A manuscript with results will be prepared and submitted for journal publication upon project completion. The findings of the study will be disseminated at a relevant (inter)national conference. The results of this research will be published in a relevant journal in the rehabilitation category, which has peer review and qualifies physical therapy research and practice. All results of this meta-research study will also be announced at (inter)national scientific events in the area of rehabilitation and research method.

## AUTHORS’ CONTRIBUTIONS

TI, AC, and RO conceived and designed the study protocol. TI, SS, SG, SM, FB and DF designed the draft search strategy. TI, SS, SG, SM, FB, DF, RO and AC were involved in conceptualising the study objectives, providing input into the search strategy, study selection criteria and plans for data extraction. All the authors, including TI, SS, SG, SM, FB, DF, RO and AC, approved the final version of the protocol.

